# Neural Anomalies during Vigilance in Schizophrenia: Diagnostic Specificity and Genetic Associations

**DOI:** 10.1101/2020.04.03.20052100

**Authors:** Samuel D. Klein, Laurie L. Shekels, Kathryn A. McGuire, Scott R. Sponheim

## Abstract

Impaired vigilance is a core cognitive deficit in schizophrenia and may serve as an endophenotype (i.e., mark genetic liability). We used a continuous performance task with perceptually degraded stimuli in schizophrenia patients (*N=48)*, bipolar disorder patients (*N=26)*, and first-degree biological relatives of schizophrenia patients (*N*=55) and bipolar disorder patients (*N* =28) and healthy controls (*N*=68) to clarify whether previously reported vigilance deficits and abnormal neural functions were indicative of genetic liability for schizophrenia as opposed to a generalized liability for severe psychopathology. We also examined variation in the Catechol-O-methyltransferase gene to evaluate whether brain responses were related to genetic variation associated with higher-order cognition. Relatives of schizophrenia patients had an increased rate of misidentification of nontarget stimuli as targets when they were perceptually similar, possibly reflecting difficulties with contour perception. Larger early visual responses (i.e., N1) were associated with better task performance, while reduced N2 augmentation to target stimuli was specific to schizophrenia patients. Both schizophrenia patients and relatives of schizophrenia patients displayed reduced late cognitive responses (P3b) that predicted worse performance. Bipolar patients and their relatives exhibited performance deficits and some aberrant neural responses that were milder and dependent on sex. Variation in the Catechol-O-methyltransferase gene was differentially associated with P3b in schizophrenia and bipolar groups. Poor vigilance in schizophrenia is specifically predicted by a failure to enhance early visual responses, weak augmentation of mid-latency brain responses to targets, and limited engagement of late cognitive responses that may be tied to genetic variation associated with prefrontal dopaminergic availability.

## 1. Introduction

Impaired vigilance represents a core cognitive deficit in schizophrenia, with evidence suggesting that it may mark genetic liability for the disorder (i.e., be an endophenotype; Cornblatt & Malhotra, 2001; Gottesman & Gould, 2003; Green et al., 2004). Patients with schizophrenia generally demonstrate poor vigilance on sustained attention tasks, with some studies reporting similarly reduced performance in first-degree biological relatives (Chen & Faraone, 2000; Demeter et. al, 2013; Nestor et al., 1990; Wohlberg & Kornetsky, 1973). The degraded stimulus continuous performance task (DS-CPT) reliably yields a linear decline in performance over the task’s duration indicative of measures of sustained visual attention (Nuechterlein et al, 1983; Nuechterlein, 1991; Parasuraman, 1979). Given that patients with schizophrenia and their biological relatives have failed to demonstrate steeper linear declines in performance compared to healthy controls—they exhibit a consistent degree of impairment in perceptual sensitivity (d′) across the duration of the task (Nuechterlein, 1983; Sponheim et al., 2006)—impaired performance may reflect underlying abnormalities in visual perception. To examine neural contributions to visual perception as tapped by the DS-CPT and to test the specificity or abnormalities to genetic liability for schizophrenia, we carried out an analysis of event-related potentials to DS-CPT stimuli in patients with schizophrenia, patients with bipolar patients, and first-degree biological relatives of both patient types.

Previous reports have documented that genetic liability for schizophrenia may be related to reduced sensitivity on CPTs that use visually degraded stimuli. In studies that employed CPTs with degraded stimuli, patients with schizophrenia had lower d′ values relative to healthy controls (Liu et al., 1997; Nuechterlein et al., 2015), with similar deficits in perceptual sensitivity observed in first-degree relatives (Asarnow et al., 2002; Grove et al., 1991; Maier et. al, 1992). Critically, the reduced sensitivity for differentiation of target stimuli from nontargets (i.e. vigilance) is not due to difficulty sustaining attention over the duration of the task since the size of the deficit is comparable at the beginning and ending blocks of trials. (Buchanan, 1997; Fleck et al., 2001; Knott et al., 1999; Nestor et al., 1990; Sponheim et al., 2006). It may be that failures in perceptual processes required to visually discriminate degraded task stimuli are the source of impaired vigilance on the DS-CPT.

The neural correlates of reduced vigilance can be evaluated using ERPs. Koelega et al. found that vigilance may be related to electrophysiological responses within the 250-650 msec after stimulus presentation (1992). In tasks tapping visual attention, a decrement in P3 components has been consistently demonstrated in schizophrenia (Martínez et al., 2012; Potts et al., 2002; VanMeerten et al., 2016; Wood et al., 2007). Specific to the DS-CPT, Knott et al. found patients with schizophrenia had a diminution in P3 amplitudes relative to healthy controls (Knott et al., 1999). Previously, we reported that early sensory responses (N1 components) were augmented in first-degree relatives by vigilance demands of the DS-CPT, but not in patients, and that later neural responses (e.g., P300) were tied to target detection and were reduced in both patients and relatives (Sponheim et al., 2006). Thus, ERPs elicited by the DS-CPT may reflect neural functions associated with genetic liability for schizophrenia (i.e., be an endophenotype).

Establishing which manifestations of psychopathology relate to an abnormality is central to establishing an endophenotype. The DS-CPT has been employed in a number of clinical populations, with evidence that the task yields specific diagnostic effects. For instance, euthymic bipolar disorder patients performed similarly to healthy controls while manic patients performed more poorly, suggesting mania impairs vigilance (Fleck et al., 2005). Given the shared genetic overlap between bipolar disorder and schizophrenia (Purcell et al., 2009) a number of studies have employed the DS-CPT in both patient types to determine if impaired vigilance is shared across forms of severe psychopathology. Results suggest that schizophrenia is differentiated from bipolar disorder by reduced perceptual sensitivity (Kumar et al., 2010; S. K. Liu et al., 2002) and longer RTs to targets (Fleck et. al, 2001). Likewise, reduced perceptual sensitivity on the DS-CPT distinguished individuals with a psychotic disorder from those with major depressive disorder (Nelson et al., 1998)). Thus, reduced perceptual sensitivity on the DS-CPT appears to differentiate schizophrenia from other forms of severe mental illness.

Identifying specific elements of genetic variation associated with a candidate endophenotype may help clarify the genetic and neurobiological substrates of the disorder (Greenwood et al., 2012). Several investigations have linked deletions of chromosome 22q11 to enhanced susceptibility of schizophrenia (Bassett et al., 1998, 1998, 2003; M. Karayiorgou et al., 2010; Mar. Karayiorgou et al., 1995). One specific deletion occurs within the region coding for the COMT gene (Brisch et al., 2009). A number of studies have related the Val^158^Met COMT single nucleotide polymorphism (SNP) to both schizophrenia and poor performance on cognitive tasks associated with frontal lobe function (Egan et al., 2001; Fan et al., 2005; Malhotra et al., 2002; Winterer et al., 2006). Critically, COMT polymorphisms implicated in schizophrenia are also associated with bipolar disorder (Shifman et al., 2004), and both disorders are characterized by impairments in higher-order cognition (Green, 2006; Morice, 1990; Vöhringer et al., 2013). COMT polymorphisms are also related to self-reported anhedonia in relatives of patients with schizophrenia, but not relatives of bipolar disorder patients (Docherty & Sponheim, 2008) suggesting differences in genetic liability for the two disorders. The COMT Val allele has been associated with positive symptoms in schizophrenia, while Met homozygosity is associated with positive symptoms in bipolar disorder (Goghari & Sponheim, 2008) also supporting a differential role in the clinical expression of the disorders. Examining how COMT gene expression alters neural responses implicated in higher-order cognition could identify unique biomarkers for schizophrenia and bipolar disorder, or elucidate shared pathophysiology between the two disorders.

In order to clarify neural abnormalities associated with vigilance deficits in schizophrenia, and to evaluate the diagnostic specificity of abnormal brain responses elicited by the DS-CPT, we included schizophrenia patients, bipolar patients, and first degree-relatives of each patient group in the current analysis. We examined a single nucleotide polymorphisms (SNPs) of Catechol-O-methyltransferase (COMT) gene to evaluate whether brain responses elicited by the DS-CPT may be specifically associated with a select aspect of genetic variation that has been related to both schizophrenia and bipolar disorder (Docherty & Sponheim, 2008; Goghari & Sponheim, 2008; Silberschmidt & Sponheim, 2008; Venables et al., 2009). The present work was designed to address three specific questions: 1) Do augmented early (i.e. N1) potentials indicative of visual processing differentiate genetic liability for schizophrenia from genetic liability for bipolar disorder?; 2). Do patients with bipolar disorder and their first-degree relatives share aberrant middle (i.e. N2) and late (i.e. P3b) latency posterior brain potentials seen in patients with schizophrenia and their relatives?; 3) Does variation in the COMT gene relate to neural functions implicated in higher-order cognition, and does this relationship differentiate genetic liability for schizophrenia from bipolar disorder?

Importantly, we contrasted event-related potentials elicited by stimuli during sensory control trials (Sponheim et al., 2006) with trials on the DS-CPT to differentiate sensory responses to the images from responses evident during the vigilance demands of the DS-CPT. We hypothesized that vigilance effects would be most evident in early neural responses associated with perceptual processing of visual stimuli in individuals with liability for schizophrenia (Pokorny et al., 2019; Schallmo et al., 2013; Silverstein et al., 2015). Likewise, we compared ERPs elicited during nontarget and target trials of the DS-CPT to test whether impaired target detection differentiated genetic liability for schizophrenia from liability for bipolar disorder. Finally, we examined associations between ERPs and performance indices of the DS-CPT with symptomatology, medication dosage, and estimated IQ to investigate relationships with aspects of disease expression.

## 2. Methods

### 2.1 Participants

**Table 1** presents participant characteristics. Stable outpatients with schizophrenia or bipolar disorder, but no history of illicit drug dependence, were recruited from the clinics of the Minneapolis Veterans Affairs (VA) Medical Center, community support programs, and county mental health clinics. We identified first-degree biological relatives by interviewing patients and inviting them to participate via letter and telephone. Nonpsychiatric controls were recruited through postings at the Minneapolis VA Medical Center and surrounding Minneapolis community, and through newsletters for veterans and fraternal organizations. Exclusion criteria for control subjects included a personal or family history of psychosis or affective disorder (DSM-IV; American Psychiatric Association, 1994) and histories of substance dependence. Subjects were not excluded for history of alcohol dependence unless they had consumed alcohol in the last month. First-degree relatives were not excluded for histories of substance dependence in order to adequately characterize families of schizophrenia probands. All participants gave informed consent, and study protocol and consent procedure were approved by both the Minneapolis VA Medical Center and University of Minnesota Institutional Review Boards. The COMT genotype of each participant was determined through a restriction fragment length polymorphism technique as described by Bergman-Jungestrom and Wingren (2001) and detailed in Venables et al. (2009). A description can also be found in the online Supplemental Materials.

**Table 1.**
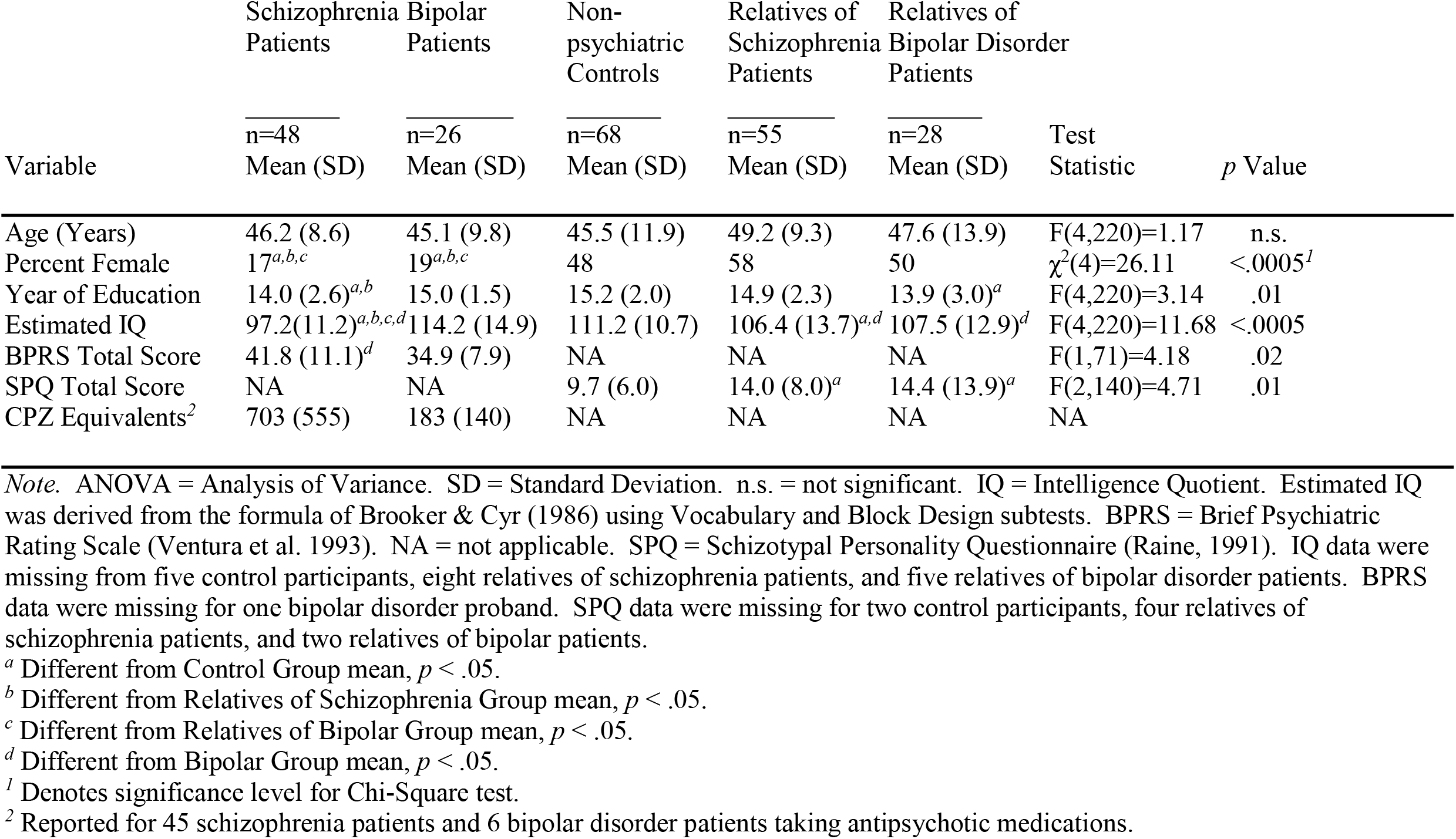
Characteristics of Participants

Trained doctoral-level clinical psychologists completed the Diagnostic Interview for Genetic Studies (DIGS; Nurnberger et al., 1994) and made symptom ratings using the Scale for the Assessment of Negative Symptoms (Andreasen, 1989), the Scale for the Assessment of Positive Symptoms (Andreasen, 1984), and the 24-item version of the Brief Psychiatric Rating Scale (Ventura et al., 2000) in order to inform clinical diagnoses. Relatives and control subjects completed the Structured Clinical Interview for DSM-IV Axis I Disorders (SCID-I; First et al., 1996). Lifetime diagnoses were determined through clinical consensus consistent with published guidelines (Leckman et al., 1982), which involved review of subjects’ clinical information from the study, requested medical history, and family informant material.

### 2.2 DS-CPT

The version of the DS-CPT employed in the present study has been described previously (Continuous Performance Test Program for IBM-Compatible Microcomputers, Version 7.10 for the Degraded Stimulus CPT, K.H. Nuechterlein and R.F. Asarnow, 1996; Sponheim at al., 2006). Briefly, task stimuli and background were degraded; 40% of white numeral pixels were switched to black, and 40% of black background pixels switched to white. Sensory control trials were administered before DS-CPT instructions were provided and consisted of “just look” (participants instructed to look passively at the screen) and “press every” (participants instructed to respond to each stimulus) at 80 trials each. Following a practice block, subjects then received DS-CPT instructions and completed three experimental blocks wherein 25% of stimuli were targets (“0”) with the remainder were nontargets (numerals “1” to “9”). Trials designated as “similar” more closely resembled targets (“6”, “8” “9”), while “dissimilar” consisted of the remaining numerals. For practice and experimental blocks, participants were told to respond only when they saw targets. Standard signal detection indices of d′ and ln(β) were computed for each subject.

### 2.3 Electrophysiological Data Collection and Analyses

EEG collection procedures for participants has been detailed previously (Sponheim et al., 2006). Electroencephalogram signals were digitized at a rate of 500 Hz with a .05-Hz low-frequency, 100-Hz high-frequency, and a 60-Hz notch filters. Recordings were divided into epochs extending from 100 msec before stimulus to 1000 msec post-stimulus. Vertical electro-oculograms were used to remove ocular artifact (Semlitsch et al. 1986) and data baseline corrected to 100 msec prior to the onset of the stimulus, and filtered with a high-frequency cutoff of 30 Hz (48 dB/octave roll-off) and a low-frequency cutoff of .1 Hz (48 dB/octave roll-off). Epochs with signals exceeding ±100 μV or horizontal electrooculogram were automatically rejected. All epochs were visually inspected, and excluded if artifacts less than 100 μV were identified. Only trials containing correct responses were averaged for control (just look, button press), target, and nontarget trials.

### 2.4 Statistical Analyses of Electrophysiology Measures

To examine how neural components associated with target detection during vigilance and sensory control trials are associated with genetic liability for schizophrenia and bipolar disorder, we conducted two sets of multivariate analyses of variance (MANOVAs) with two between-subjects factors (group and gender) for each ERP component of interest (i.e. N1, N2, P3b). In one analysis set, the group factor consisted of healthy controls (HC), patients with schizophrenia (SZ) and first-degree relatives of patients with schizophrenia (SZRel), while the second set consisted of HC, bipolar probands (BP) and first-degree relatives of bipolar patients (BPRel). For analyses relating to N1, three within-subject factors were used (target: target vs. nontarget, hemisphere: left vs. right, task: vigilance vs. sensory control trials) because the component was evident across all task conditions. We computed the N2 difference wave as the amplitude to nontarget objects subtracted from target amplitude because it was only evident during the DS-CPT trials and was most relevant to cognitive control processes related to target identification (Swainson et al., 2003). Only one within-subjects factor (region: electrode Cz vs. electrode Fz) was used for these analyses. Finally, because P3b was only evident to target objects, the within-subjects factor of hemisphere and task (vigilance vs. “press only” trials) was exclusively used to examine how the vigilance and target identification demands of the DS-CPT affected the component beyond the motor response demands of both trial types. In order to account for the shared genetic contribution present amongst individual families biasing our results, we performed mixed effects models with family membership included as a random effect (see online Supplementary Table S1).

## 3. Results

### 3.1 Task Performance

Behavioral results as well as contrasts involving all groups and participants are displayed in **Table 2**. A closer examination of the distribution of d′ scores revealed 3 exceptionally high-performing outliers among SZ, and one among SZRel according to procedures outlined in Schwertman et al. (2004). There were no other behavioral outliers in any other group. Planned comparisons between HC and SZ after removing said outliers revealed SZ performed worse than HC on block 2 (FDR corrected *p* = .024) and overall (FDR corrected *p* = .044), but did not change group differences between HC and SZRel (a comparison of participant characteristics between SZ in the present sample, and SZ from the sample in our previous report can be found in Supplementary Table S2). In order to account for the unequal distribution of male and female subjects in proband groups and to test for whether performance deficits were specific to liability for a particular disorder, or indicative of liability for severe mental illness in general, two 3 × 2 Mixed Model ANOVAs were fitted with between-subjects factors of group and gender, and a within-subjects factor of experimental block (first, second, and third). The first ANOVA tested differences between HC, SZ and SZRel. There was a main effect of block (F_(*2, 161)*_ = 56.09, p < .001, η^2^ = .258), with average performance decreasing on later blocks, with no effect of group (F_(*2, 161)*_ = 2.05 p = .132, η^2^ = .025), gender (F_(*1, 165)*_ = .190, p = .542, η^2^ = .001) and no observed interaction between group and gender (F_(*2, 165)*_ = .717, p = .74, η^2^ = .004). The second ANOVA examined differences between HC, BP and BPRel. There was a main effect of block (F_(*2, 116)*_ = 17.21, p < .001, η^2^ = .129), group (F_(*2, 116)*_ = 5.41, p < .01, η^2^ = .086), with no effect of gender (F(_*1, 116)*_ = 2.39, p = .125, η^2^ = .020) and an interaction between group and gender (F_(*2, 116)*_ = 3.12, p =.048, η^2^ = .051), as well as an interaction between block, group and gender (F_(*2, 116)*_ = 4.00, p =.014, η^2^ = .052). Simple main effects revealed that performance deficits were specific to male BP (M = 2.21, SE = .212; Sidak corrected *p =* .05) and male BPRel (M = 2.00, SE = .259; Sidak corrected *p =* .018) compared to male HC (M = 2.86, SE = .164), with the greatest differences occurring during blocks 1 and 2 of the task. Across all subjects, total d′ was associated with IQ (*r* (207) = .31, FDR corrected *p <* .01), and this association was also seen in BP (*r*(26) = .60, FDR corrected *p* = .02) as well SZRel (*r* (47) = .55, FDR corrected *p* < .01).

**Table 2.** Degraded Stimulus Continuous Performance Task (DS-CPT) Performance

| Task Value<sup>1</sup> | Schizophrenia Patients<br>n=48<br>Mean (SD) | Bipolar Patients<br>n=26<br>Mean (SD) | Non-psychiatric Controls<br>n=68<br>Mean (SD) | Relatives of Schizophrenia Patients<br>n=55<br>Mean (SD) | Relatives of Bipolar Patients<br>n=28<br>Mean (SD) | ANOVA Test Value | Effect Size-<br>$\eta^2$ | *p* |
| --- | --- | --- | --- | --- | --- | --- | --- | --- |
| Target Detection: *d'* |  |  |  |  |  |  |  |  |
| Block 1 | 2.67 (.89) | 2.67 (1.08) | 3.02 (1.06) | 2.85 (.89) | 2.23 (1.20)<sup>a,b</sup> | F(4,220)=3.25 | .046 | .01 |
| Block 2 | 2.38 (.96) | 2.40 (1.14) | 2.75 (1.10) | 2.59 (.86) | 1.97 (1.00)<sup>a,b</sup> | F(4,220)=3.31 | .047 | .01 |
| Block 3 | 2.27 (.96) | 2.21 (.93) | 2.49 (1.08) | 2.56 (.88) | 1.96 (.87)<sup>a,b</sup> | F(4,220)=2.37 | .041 | .05 |
| Total | 2.42 (.93) | 2.37 (.98) | 2.73 (1.10) | 2.64 (.89) | 1.99 (.89)<sup>a,b</sup> | F(4,220)=3.37 | .05 | .01 |
| Similar-Dissimilar False Alarms | 12.89 (9.6) | 11.69(10.5) | 9.70(10.9) | 16.03(10.2)<sup>a</sup> | 10.3(12.4) | F(4,220)=2.81 | .049 | .03 |
| ln-β | | | | | | | | |
| Total | .69 (.79) | .98 (.85) | .68 (.85) | .68 (1.0) | 1.0 (.69) | F(4,220)=1.9 | .021 | .31 |
| Reaction Time to Targets: (msec) |  |  |  |  |  |  |  |  |
| Total | 529 (88)<sup>a,b</sup> | 531 (68)<sup>a,b</sup> | 481 (61) | 493 (55) | 548 (95)<sup>a,b</sup> | F(4,220)=6.75 | .109 | <.001 |

Note. ANOVA = Analysis of Variance. SD = Standard Deviation. n.s. = not significant.
<sup>1</sup>Denotes significance level of One-way ANOVA
<sup>a</sup> Different from Control Group mean, Tukey's HSD $p \leq .05$
<sup>b</sup> Different from Relatives of Schizophrenia Group mean, Tukey's HSD $p \leq .05$

DS-CPT performance indices failed to be associated with symptom measures in patients and schizotypy scales (SPQ) in relatives.

To examine the hypothesis that perceptual processes related to contour detection affect performance in individuals with genetic liability for schizophrenia, we carried out exploratory analyses of the rate of incorrect target identifications for nontargets that shared curved contours with the target ‘0’ (numerals ‘6’, ‘8’ and ‘9’). Two 3 × 2 ANOVAs with between subjects factors of group and gender were employed to examine the rate of false alarmsto similar nontargets while subtracting errors for dissimilar nontargets to control for overall false alarm rate (i.e., SimDiff Errors; **Table 2)**. The first ANOVA examining healthy controls, patients with schizophrenia and their first-degree relatives yielded a main effect of group (F_(*2, 167)*_ = 3.19, p = .01, η^2^ = .054). Follow up post-hoc tests revealed SZRel had greater SimDiff errors than HC (Tukey’s HSD *p <* .01) while the errors for SZ failed to be significantly higher (Tukey’s HSD p=.379). A separate 3 × 2 ANOVA examining healthy controls, BP and BPRel yielded a significant effect of gender (F_(1, 116)_ = 3.19, p = .047, η^2^ = .034) and an interaction between group and gender (F_(2, 116)_ = 3.36, p = .038, η^2^ = .055). Follow up simple main effects tests revealed that female BPRel (M=16.07, SE = 2.60) had substantially more SimDiff errors than male BPRel (M = 4.5, SE = 2.60; Sidak corrected *p* < .01).

### 3.2 ERP Results

#### 3.2.1 Neural Anomalies and Genetic Liability for Schizophrenia

##### Early Posterior Potential: N1

A MANOVA examining the N1 component in HC, SZ and SZRel at O1 and O2 revealed a main effect of task (*F*_(*1,161)*_ = 19.60, *p* <.001, Wilk’s Λ =.891, partial η^2^ = .109), where N1 was greater during vigilance (M = -5.42, SE =.42) than sensory control trials (M = -4.43, SE = .394; Sidak corrected *p* < .001). There was also an interaction between task, group and gender (*F*_(*2,161)*_ = 4.25, *p* = .016, Wilk’s Λ =.95, partial η^2^ = .05). Follow up simple main-effects analyses revealed that N1 amplitudes were greater to target (Sidak corrected *p* < .001) and nontarget trials (Sidak corrected *p* = .032) during vigilance than for sensory control trials, particularly at electrode site O2 (Sidak corrected *p* < .001). There were trends for effects of gender (F_(1,161)_ = 3.38, p = .068, η^2^ = .024) and group (F_(2,161)_ = 2.54, p = .082, η^2^ = .031), but no interaction between group and gender (F_(2,161)_ = .762, p = .468, η^2^ = .009). Planned comparisons revealed that SZRel had greater N1 amplitudes than both HC (FDR corrected *p* = .03) and SZ (FDR corrected *p* = .02.) with this difference being greatest at site O2 for targets during vigilance **(Figure 1A)**. Additional follow-up simple main effects analyses revealed that female SZRel (M = -7.81, SE = .833) differed from female HC during sensory control trials (M = -5.84, SE = .83; Sidak corrected *p =* .02).

**Figure 1.**
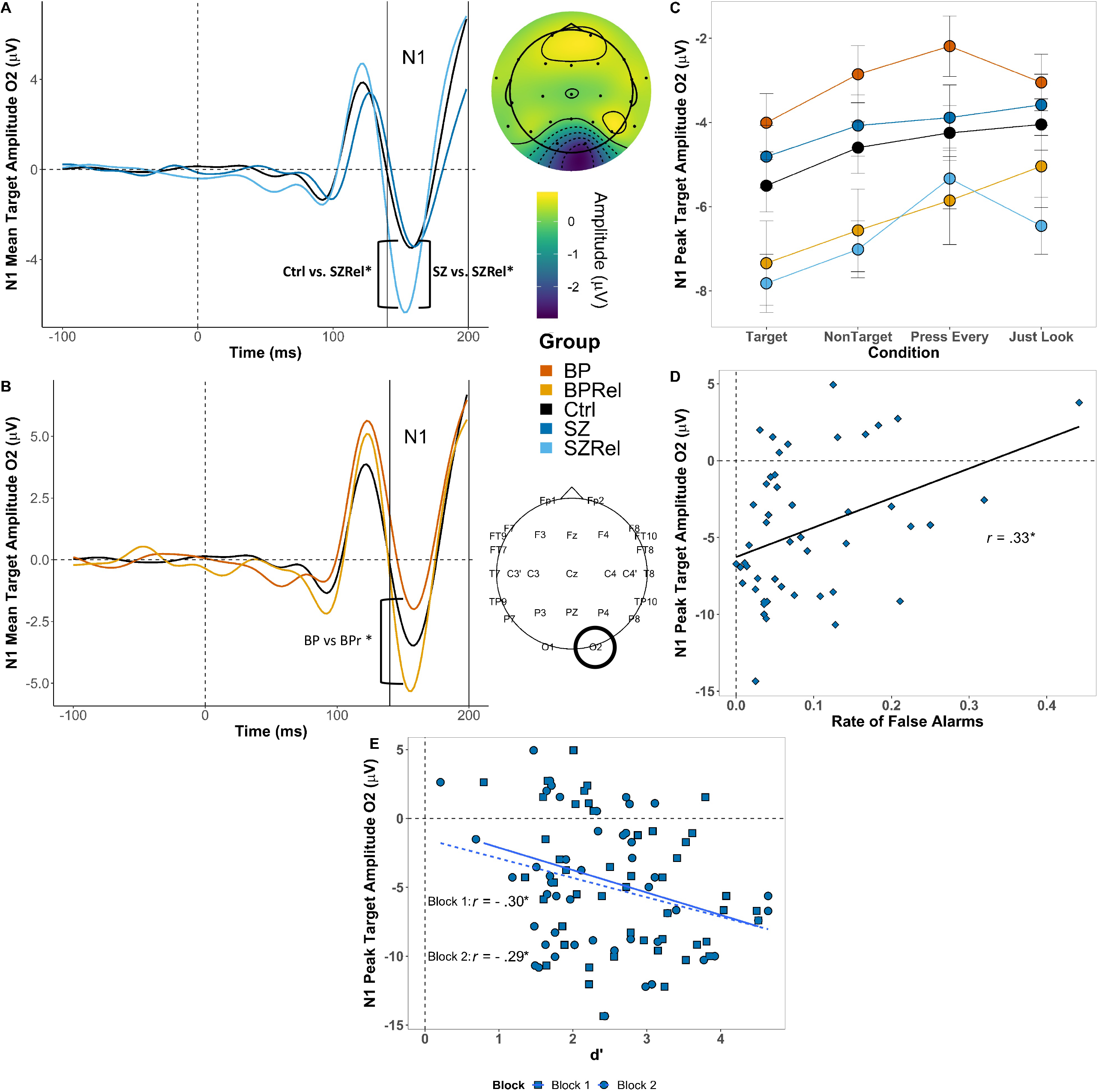
Grand averaged N1 to targets at O2. (A) SZRel had greater peak amplitude compared to HC (FDR corrected *p* = .035) and SZ (FDR corrected *p* = .015. (B) BPRel had greater peak amplitude compared to BP (FDR corrected *p* = .014) but not HC (FDR corrected *p* = .119). (C) N1 amplitudes across groups and conditions at O2 (error bars ± SEM). (D and E) N1 amplitude to targets at O2 was positively associated with the number of false alarms (*r(48) =* .33, FDR corrected *p* = .04; D) and d′ in block 1 (*r*(48) *=* -.30, FDR corrected *p* = .04) and block 2 (*r*(48) *= -*.29, FDR corrected *p* = .05; E) in SZ. * indicates p < .05 | ** indicates *p* < .01

The effect of N1 across conditions is presented in **Figure 1C**. A main effect of hemisphere was also observed (*F*_(*1,161)*_ = 6.77, *p* = .01, Wilk’s Λ =.987, partial η^2^ = .04) such that amplitudes were greater at O2 (i.e. right hemisphere; M = -5.17, SE = .42) than at O1 (M = -4.68, SE =.39; Sidak corrected *p* = .01). There were interactions between task and target (*F*_(*1,161)*_ = 5.48, *p* = .02, Wilk’s Λ =.967, partial η^2^ = .03) and between task and hemisphere (*F*_(*1,161)*_ = 4.02, *p* = .047, Wilk’s Λ =.976, partial η^2^ = .024).

Correlations were computed between N1 amplitude to targets at site O2 and behavioral indices on the DS-CPT. In SZ, N1 amplitude was positively associated with the false alarm rate (*r(48) =* .33, FDR corrected *p* = .04; **Figure 1D**), suggesting that *reduced* augmentation of early posterior brain responses to targets contributed to a tendency to identify nontarget stimuli as targets. N1 amplitudes were additionally negatively correlated with d′ during block 1 (*r*(48) *=* -.30, FDR corrected *p* = .04) and block 2 (*r*(48) *= -*.29, FDR corrected *p* = .05) in SZ also suggesting that robust early posterior responses to stimuli yielded better differentiation of target and nontarget stimuli (**Figure 1E)**.

##### Middle Latency Potential: N2

A MANOVA examining group differences between HC, SZ and SZRel revealed a main effect of group (F_(2,161)_ = 5.30, p < .01, η^2^ = .061) with follow-up post-hocs revealing that SZ had reduced amplitudes compared to HC (Tukey HSD *p* <.01), with this difference being greatest at site Cz to targets during vigilance **(Figure 2A)**. There was a trend towards a gender effect (F_(1,161)_ = 2.85, p < .093, η^2^ = .017), and no interaction between gender and group (F_(2,161)_ = .251, p =.778, η^2^ = .003). There were also interactions between electrode site and group (*F*_(*2,162)*_ = 5.33, *p* <.01, Wilk’s Λ =.938, partial η^2^ = .062) and electrode site and gender (*F*_(*1,161)*_ = 4.19, *p* = .042, Wilk’s Λ =.938, partial η^2^ = .025). An effect of electrode (*F*_(*1,1621)*_ = 61.42, *p* <.001, Wilk’s Λ =.725, partial η^2^ = .275) reflected that N2 difference waveforms were greater at Cz (M = -8.14, SE = .308) than Fz (M = .595, SE = .246; Sidak corrected *p <* .001). Follow up analyses for each electrode site revealed that SZ had reduced N2 difference waveforms at Cz (M = 1.307, SE = .534) compared to HC (M = -.240, SE = .336; Sidak corrected *p* = .045) and reduced N2 difference waveforms at Fz (M = .831, SE = .67) compared to both HC (M = -2.00, SE = .422; Sidak corrected *p* < .01) and SZRel (M = -1.28, SE = .48; Sidak corrected *p* = .035). Further simple main effects analyses showed that females had greater difference waveforms at Cz (M =-.185, SE =.497) than males (M =.857, SE =.396; Sidak corrected *p* = .045).

**Figure 2.**
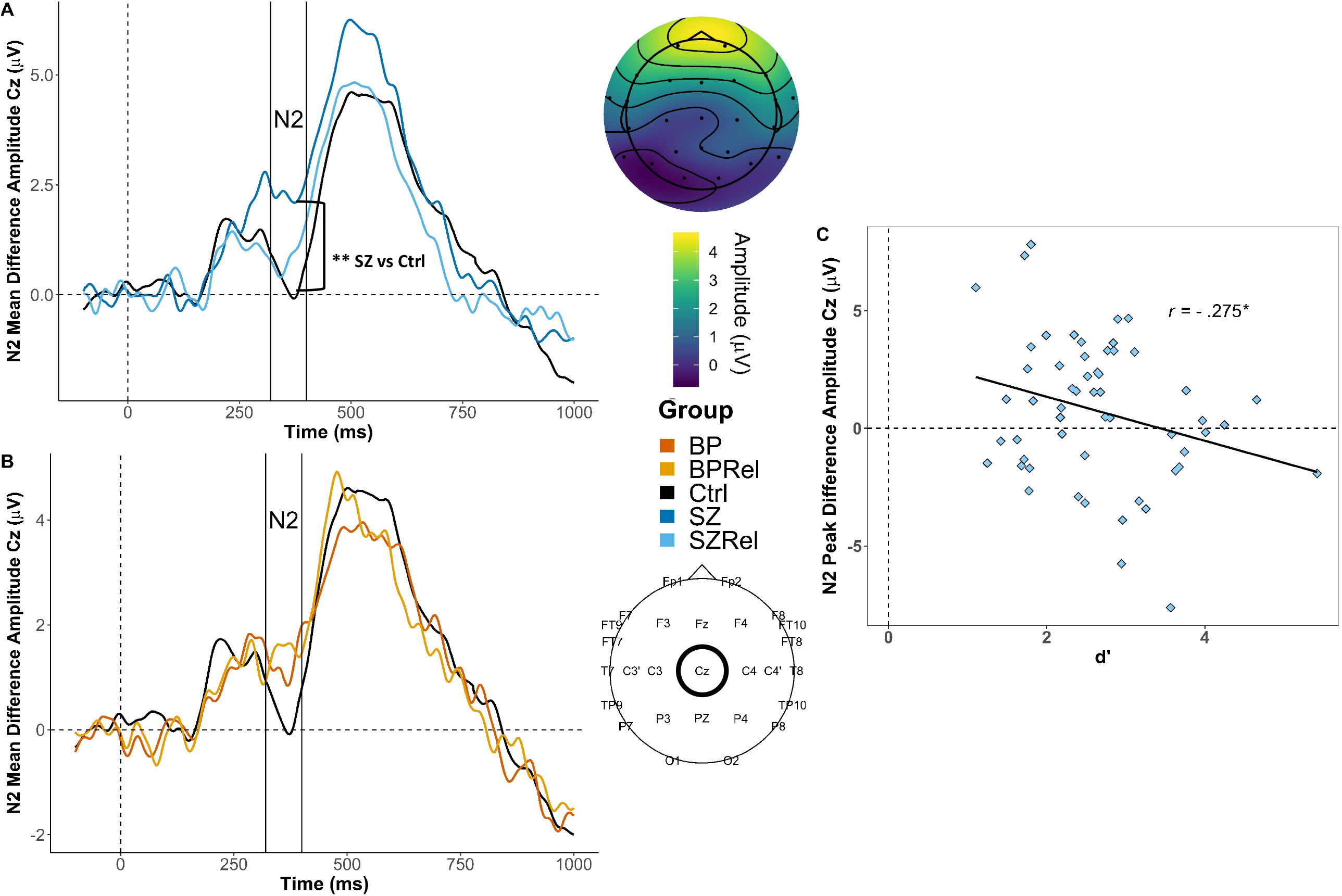
Grand averaged N2 difference waveforms (targets – nontargets) at Cz. (A) SZ had reduced peak amplitudes compared to HC (FDR corrected *p* < .01) (B) There were no observed group differences in peak N2 difference waveforms between BP, BPRel and HC (C) N2 amplitudes were positively associated with average d′ in SZRel (*r(54) =* -.275, FDR corrected *p* = .05). * indicates p < .05 | ** indicates *p* < .01

Correlations were computed between difference waves at site Cz and performance indices on the DS-CPT, as well as IQ and clinical symptom ratings. In SZRel, there was a significant association between total d′ (across all three blocks) and N2 (*r(54) =* -.275, FDR corrected *p* = .05), such that larger N2 responses to target stimuli were associated with better differentiation between target and nontarget stimuli on the DS-CPT (**Figure 2C)**.

##### Late Posterior Potential: P3b

A MANOVA examining the P3b component to targets at electrode sites P7 and P8 in HC, SZ and SZRel revealed a significant effect of task (*F*_(*1,162)*_ = 167.02, *p* <.001, Wilk’s Λ =.492, partial η^2^ = .508) where amplitudes were greater during vigilance (M = 7.23, SE =.29) relative to “press every” control trials (M =3.12, SE = .178; Sidak corrected *p* < .001). There were also main effects of group (*F*_*(2,162)*_ = 3.14, *p* =.016, partial η^2^ = .05), gender (*F*_*(1,162)*_ = 6.031, *p* =.015, partial η^2^ = .036), and an interaction between gender and group (*F*_*(2,162)*_ = 3.161, *p* =.045, partial η^2^ = .038). Planned comparisons revealed that SZ and SZRel had smaller P3bs than HC (FDR corrected *p <* .01; FDR corrected *p =* .023 respectively), particularly at electrode site P7 for target stimuli during vigilance (**Figure 3A)**. There was an interaction between hemisphere and group (*F*_(*2,162)*_ = 3.14, *p* =.046, Wilk’s Λ =.963, partial η^2^ = .204), with SZ (M =4.43, SE =.413) having reduced P3b components compared to HC in the left hemisphere (i.e. P7; M =5.92, SE =.262; Sidak corrected *p* <.01) and SZRel (M =4.94, SE = .295) also displaying reduced P3b compared to HC at P7 (Sidak corrected *p =* .042). Follow-up simple main effects revealed that gender differences were only apparent in HC (*F*_*(1,162)*_ = 11.454, *p* <.041, partial η^2^ = .066), with females having greater P3b compared to males (Sidak corrected *p* < .01). The only association specific to schizophrenia was that larger P3b amplitude at electrode site P7 was associated with higher daily dosages for antipsychotic medication (chlorpromazine equivalence; *r*(45) *=* .412, FDR corrected *p <* .01).

**Figure 3.**
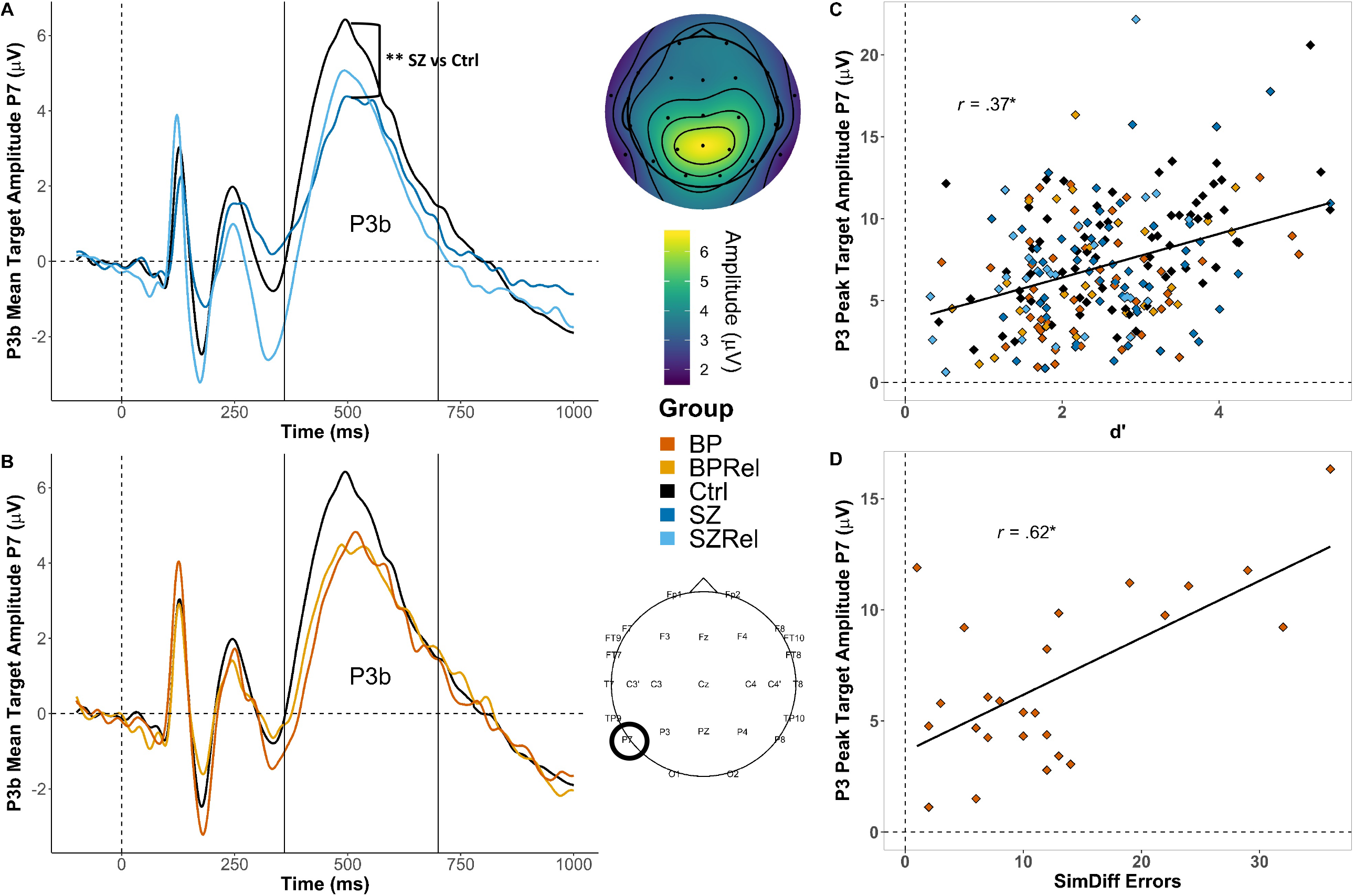
Grand averaged P3b amplitude to targets at P7. (A) SZ had reduced peak amplitudes compared to HC (FDR corrected *p* < .01) (B) There were no observed group differences in peak P3b amplitudes between BP, BPRel and HC at P7 alone (C) P3b amplitudes were positively associated with average d′ across all five groups (*r*(225) = .37, FDR corrected *p <* .01) D) P3b amplitudes were positively associated with the number of SimDiff errors in BP (*r* (25) = .619, FDR corrected *p* <.01). * indicates p < .05 | ** indicates *p* < .01

#### 3.2.2 Neural Anomalies and Genetic Liability for Bipolar

##### Disorder Early Posterior Potential: N1

A MANOVA examining N1 amplitudes at O1 and O2 in HC, BR and BPRel revealed a main effect of task (*F*_(*1,111)*_ = 10.67, *p* <.01, Wilk’s Λ =.912, partial η^2^ = .088), with N1 responses being greater during vigilance (M = -4.84, SE = .50) than sensory control trials (M = -3.77, SE = .329; Sidak corrected *p* < .01). There was also an effect of group (*F*_*(2,111)*_ = 3.31, *p* =.04, partial η^2^ = .056), with follow-up post hoc tests revealing that BPRel had greater N1 amplitudes than BP (Tukey’s HSD, *p* = .011). Group differences were maximal at electrode O2 to targets during vigilance (**Figure 1B)**. Correlations were computed between N1 amplitude to targets and behavioral indices on the DS-CPT in BP and BPRel with no correlations surviving FDR correction.

##### Middle Latency Potential: N2

A MANOVA examining N2 difference waveforms at electrode sites Cz and Fz revealed an effect of region (*F*_(*1,115)*_ = 49.55, *p* <.001, Wilk’s Λ =.70, partial η^2^ = .301), with N2 amplitudes being greater at Cz (M = -1.18, SE = .33) than Fz (M =.405, SE =.326; Sidak adjusted *p* < .001). There was no effect of group (*F*_*(2,115)*_ = 1.96, *p* =.145, partial η^2^ = .033), gender (*F*_*(1,115)*_ = 2.73, *p* = .101, partial η^2^ = .023) and no interaction between group and gender (*F*_*(1,115)*_ = .524, *p* = .593, partial η^2^ = .009). Planned comparisons revealed no group differences after FDR correction. Correlations were computed between difference waves at site Cz and performance indices on the DS-CPT, as well as IQ and clinical symptom ratings. No correlations in patients with bipolar disorder or their first-degree relatives survived FDR correction.

##### Late Posterior Potential: P3b

A MANOVA examining P3b amplitudes to targets at electrode sites P7 and P8 compared HC, BP and BPRel. There was a main effect of task (*F*_(*1,112)*_ = 70.65, *p* <.001, Wilk’s Λ =.613, partial η^2^ = .387), with P3b amplitudes being greater during vigilance (M = 7.08, SE =.35) than “press every” trials (M = 3.57, SE = 2.54; Sidak corrected *p* <.001). There were also interactions between hemisphere and gender (*F*_(*1,112)*_ = 5.54, *p* = .02, Wilk’s Λ =.953, partial η^2^ = .047) and hemisphere and task (*F*_(*1,112)*_ = 11.92, *p* <.01, Wilk’s Λ =.904, partial η^2^ = .096). There were also main effects of group (*F*_*(2,112)*_ = 4.137, *p* = .018, partial η^2^ = .060), gender (*F*_*(1,112)*_ = 6.28, *p* = .014, partial η^2^ = .053) and an interaction between gender and group (*F*_*(2,112)*_ = 3.31, *p* = .04, partial η^2^ = .056). Follow-up analyses revealed that female BPRel had reduced P3b amplitudes compared to both HC (Sidak corrected *p* < .01) and BP (Sidak corrected *p* = .048). Planned comparisons revealed HC had greater P3b amplitudes compared to BPRel (FDR corrected *p* = .015) with a trend towards being greater than BP (FDR corrected *p* = .071), with the greatest differences observed at electrode site P7 during vigilance (**Figure 3B)**. There was also an interaction between task, side and group (*F*_(*2,112)*_ = 3.55, *p* = .032; Wilk’s Λ =.94, partial η^2^ = .06) with follow up simple main effects analyses revealing BPRel had smaller P3b amplitudes at both parietal electrodes (site P7 - BPRel: M =6.27, SE =.622; HC: M= 8.15 SE =.387; Sidak corrected *p* = .034; site P8 – BPRel: M =5.64, SE =.64; HC: M = 7.94, SE =.399; Sidak corrected *p* < .01). Additional follow-up analyses revealed females had greater P3b amplitudes in the left hemisphere (i.e. P7; M =6.09, SE = .38) compared to males (M = 4.58, SE = .267; Sidak corrected *p* < .01), and that during vigilance subjects had greater P3b amplitudes at P7 (M = 7.32, SE = .36) than P8 (M= 6.83, SE =.37; Sidak corrected *p* = .018). In contrast, subjects had greater P3b amplitudes during “press every” trials at P8 (M =3.35, SE = .275) than P7 (M= 3.35, SE =.275; Sidak corrected *p <* .045).

Correlations between P3b amplitude to targets at electrode P7 and behavioral indices on the DS-CPT and demographics were computed. Across all groups (i.e.HC, SZ, SZRel, BP and BPRel) larger P3b amplitudes were related to better target stimulus detection on the DS-CPT (d′: *r*(225) = .37, FDR corrected *p <* .01; **Figure 3D**) and estimated IQ (*r*(207) *=* .25, FDR corrected *p* < .01). The association between with P3b amplitude and performance was also evident in HC (d′: *r*(68) *=*.514, FDR corrected *p* <.01), as was fewer misidentifications of nontarget stimuli as targets (*r*(68) *=* -.368, FDR corrected *p* < .01). Interestingly, in BP P3b amplitudes were associated with a tendency to confuse similar nontarget stimuli with targets (SimDiff errors: *r* (25) = .619, FDR corrected *p* <.01; **Figure 3D)** possibly suggesting that the late ERP for this group reflected the perception that the nontarget stimulus was a target.

### 3.3 COMT Genotype and ERP Results

Given that COMT polymorphisms are generally implicated in prefrontal cortical networks involved in cognitive control and working memory processes (Cools & D’Esposito, 2011) we focused on the associations between COMT with ERP components previously associated with cognitive processes (N2 and P3b). To further restrict analyses, we only examined the associations between COMT and N2 in relation to schizophrenia given the lack of group differences in the bipolar disorder-related samples. Additionally, there were no female SZ that were met homozygotes and only a single female BP that was a val/met heterozygote which precluded the inclusion of gender as a factor in statistical analyses of COMT variation (i.e., 3 X 2 Mixed Model MANOVAS with the factors of group and genotype were used).

A MANOVA examining P3b effects at P7 and P8 among HC, SZ and SZRel included a genotype distribution of val homozygotes (*N =*34), val/met heterozygotes (*N=* 61) and met homozygotes (*N=*28). There was a main effect of genotype (*F*_(*1,114)*_ = 3.37, *p* =.035;, partial η^2^ = .056), but no effect of diagnostic group (*F*_(*2,114)*_ = 2.05, *p* =.134, partial η2 = .035), and no interaction between diagnostic group and genotype (*F*_(*2,114)*_ = 1.12, *p* =.352, partial η^2^ = .038). The analysis also revealed an effect of task (*F*_(*1,114)*_ = 133.88, *p* <.001, Wilk’s Λ =.46, partial η^2^ = .54), with P3 amplitudes being greater during vigilance (M = 6.81, SE = .33) than “press every” trials (M =3.13, SE =.17; Sidak corrected *p* < .001) and an interaction between task and genotype (*F*_(*2,114)*_ = 4.11, *p* =.019, Wilk’s Λ =.46, partial η^2^ = .54) with val homozygotes (M = 5.42, SE=.60) having reduced P3b amplitudes during vigilance compared to val/met heterozygotes(M = 7.01, SE = .43; Sidak corrected p =.034) and met homozygotes (M = 8.02, SE =.64; Sidak corrected *p* <.01)

A MANOVA examining P3b effects at P7 and P8 among HC, BP and BPRel included a genotype distribution of val homozygotes (*N =*17), val/met heterozygotes (*N=* 40) and met homozygotes (*N=*20). There were no main effects of group (*F*_(*2,68)*_ = 1.012, *p* =.36; partial η^2^ = .03) nor genotype group (*F*_(*2,68)*_ = 1.007, *p* =.37; partial η^2^ = .03), but there was a trend for an interaction between diagnostic group and genotype (*F*_(*2,68)*_ = 2.48, *p* =.051; partial η^2^ = .128). There was an effect of task condition (*F*_(*1,68)*_ = 90.24, *p* <.001; Wilk’s Λ =.43, partial η^2^ = .57) with P3b amplitudes being greater during vigilance (M = 6.80, SE = .35) than “press every” trials (M =3.13, SE =.25; Sidak corrected *p* < .001) and an interaction between task and genotype (*F*_(*1,68)*_ = 5.47, *p* <.01;Wilk’s Λ =.861, partial η^2^ = .14). Follow up simple main effects tests revealed that met homozygotes (M = 8.04, SE=.64) had greater P3b amplitudes than val/met heterozygotes (M = 6.01, SE = .45; Sidak corrected *p =*.03). There was also an interaction between hemisphere and task (*F*_(*1,68)*_ = 6.97, *p* <.001; Wilk’s Λ =.91, partial η^2^ = .093), but follow up simple main effects analyses did not survive correction for multiple comparisons.

To further understand the relationship between P3b amplitude and the COMT genotype we examined associations between neural functions and behavioral and demographic indices for each genotype within samples related to schizophrenia and bipolar disorder. For val homozygotes with liability for schizophrenia, total d′ across blocks was positively associated with P3b amplitude to targets at site P7 (*r(*28) *=* .456, FDR corrected *p* = .04). In individuals with liability for bipolar disorder correlations between DS-CPT performance and neural functions did not survive FDR correction. To examine the effects of medication on P3b, we examined these associations across genotype groups. Chlorpromazine equivalence was positively associated with P3b in schizophrenia probands that were val homozygotes (*r*(15) = .622, FDR corrected *p* = .04). There were no associations between P3b and medication in bipolar probands. Analyses of N2 failed to yield any effects involving genotype (see Supplementary Materials).

### 3.4 Discussion

In this study we examined behavioral performance and neurophysiological responses to the DS-CPT in healthy controls, patients with schizophrenia, patients with bipolar disorder, and first-degree biological relatives of patients with both disorders to determine whether previously reported abnormalities in brain responses were associated with genetic liability for bipolar disorder as well as schizophrenia. First-degree relatives of both patient types exhibited augmented N1 components compared to respective patient groups suggesting possible early compensatory visual functions during vigilance for visual targets that are perceptually difficult to discern. Diminished fronto-central N2 difference waveforms were specific to schizophrenia patients and may reflect disorder-specific deficits in high-level object recognition (Luck et al., 2009) or sensitivity to the degree of perceptual deviation of targets from nontargets (Folstein & Petten, 2008). Both patients with schizophrenia and their relatives exhibited reduced P3b components compared to controls and therefore the component may tap aspects of genetic liability for schizophrenia (i.e., be an endophenotype). Neurophysiological responses of cognitive control (P3b) were associated with COMT gene variation. COMT val homozygotes had the smallest P3b amplitudes when examining liability for schizophrenia, whereas val/met heterozygotes had the smallest P3b amplitudes with respect to liability for bipolar disorder, suggesting COMT variation may differentially influence neural functions indicative of higher-order cognition across the two severe mental disorders.

Relative groups of both types exhibited augmented N1 components across vigilance and sensory control conditions in contrast to comparable patient groups, though only relatives of patients with schizophrenia had larger N1s than healthy controls. In relatives of patients with schizophrenia, augmented N1 appears to be a compensatory component given 1) relatives demonstrated intact performance on the DS-CPT and 2) augmented N1 was associated with better performance in patients with schizophrenia suggesting a beneficial role of early visual cortical responses in discriminating degraded visual stimuli. This finding is consistent with a recent report in which augmented ERPs at 200 ms (comparable to N1) to target verniers in a backward masking task reflected neural compensation in relatives of patients with schizophrenia (da Cruz et al., 2020). In contrast, we do not interpret N1 as compensatory in the context of relatives of bipolar relatives given the lack of observed associations between N1 and performance in both patients with bipolar disorder or first-degree relatives, and that relatives’ N1 potentials did not differ from healthy controls.

Results also suggest that diminished N2 difference waveforms are specific to patients with schizophrenia. Previous work has demonstrated that reductions in N2 potentials reflect a broad attentional deficit in schizophrenia (Salisbury et al., 1994), with deficits in N2 difference waveforms related to a dysfunction in classification of stimuli as targets during stimulus identification (Wood et al., 2006). Consistent with a target identification function, larger N2 difference waveforms in relatives of patients with schizophrenia were associated with greater d′, suggesting that neural responses differentiating targets and nontarget stimuli facilitate DS-CPT behavioral performance. Importantly, the present findings indicate that deficits in fronto-central N2 responses reflect deficits in neural functions related to visual object recognition that are specific to schizophrenia pathology (Doniger et al., 2002).

The present study replicated previous findings of impaired P3b in both patients with schizophrenia and their relatives suggesting deficits in neural functions implicated in target detection may constitute an endophenotype of schizophrenia (Groom et al., 2008; Knott et al., 1999; Roxborough et al., 1993; Sponheim et al., 2006). In contrast, deficits in P3b were specific to relatives of patients with bipolar disorder. Critically, the observed interaction between gender and diagnostic group in models examining specificity for schizophrenia appeared to be driven by sex differences in healthy controls, consistent with hypotheses that females have larger P3b components compared with males (Conroy & Polich, 2007; Steffensen et al., 2008). This same interaction in models examining the specificity of bipolar disorder revealed that female relatives had reduced P3b components compared to both healthy controls and bipolar probands. Results from the present work suggest abnormalities in fronto-posterior parietal networks constitute an endophenotype for schizophrenia, and may be indicative of neural expression of genetic liability for bipolar disorder in female relatives.

Variation in the COMT genotype failed to be strongly related to differences in neural responses between the clinical disorders of schizophrenia and bipolar disorder. However, when examining genetic liability for schizophrenia, val/val homozygotes had the most pronounced deficits in P3b amplitudes, consistent with findings that increased dopaminergic catabolism in prefrontal neurons impairs higher order cognition in schizophrenia (Bilder et al., 2002; Roffman et al., 2008; Shifman et al., 2002). In contrast, when examining genetic liability for bipolar disorder, val/met heterozygotes had the smaller P3b amplitudes relative to met/met homozygotes. These results align somewhat with previous report in which val and met homozygotes had differential associations with clinical symptomatology in schizophrenia and bipolar disorder respectively (Goghari & Sponheim, 2008). Given that met homozygotes generally outperform the other genotypes in tasks tapping higher-order processing (Bruder et al., 2005; Tsai et al., 2003), our present findings suggest higher order cognition may be differentially impacted by COMT polymorphisms across liability for schizophrenia and bipolar disorder.

Contrary to expectations, we did not observe diminished perceptual sensitivity in patients with schizophrenia. In the new sample, d′ was more similar to what would be expected in healthy controls (Nuechterlein et al., 2015), and was significantly greater than we observed in another sample from our laboratory. Patients with schizophrenia in the present sample had a larger daily dose of chlorpromazine equivalence. Given the positive association between P3b amplitudes to targets and chlorpromazine equivalence in the present sample, higher doses of medication may have improved neural functions associated with target detection (Sponheim et al., 2006), indicating that medication effects in the present sample may mark effective treatment rather than disease severity (Nuechterlein et al., 2015). Earle-Boyer et al. demonstrated that unmedicated patients made more errors than medicated patients when performing various CPTs regardless of stimulus modality (1991), and a meta-analysis has provided evidence that antipsychotic medication mitigated P300 deficits in patients with schizophrenia (Bramon, 2004). Critically, when three outliers for performance were considered, patients with schizophrenia performed more poorly than healthy controls.

Our findings of a male specific decrement in d′ in bipolar probands and their relatives is consistent with previous reports documenting reduced attention and visual discrimination in male bipolar patients (Barrett et al., 2008; Gogos et al., 2010), and that vigilance deficits may constitute an endophenotype of bipolar disorder (Bora et al., 2009). Given that relatives of bipolar patients showed signs that they exhibited some mild symptomatology—evidenced by elevations in SPQ scores—greater reaction times in this group may reflect impaired concept shifting (Arts et al., 2008). Likewise, relatives of patients with schizophrenia showed similar signs of subtle symptomatology. In this context, greater SimDiff errors in these relatives partially supports the hypothesis that impaired contour detection is associated with genetic liability for schizophrenia.

The present study has limitations that warrant consideration. The imbalance between number of male and female probands complicates the observed interactions between diagnostic group and gender: unequal sample sizes across categorical subgroups inflates the type I error rate (Aguinis et al., 1999). Given that all observed statistical interactions between diagnostic group and gender in the present study have a smaller partial η^2^ than the main effect of group alone suggests that diagnostic group differences appear to better account for observed differences in behavior and neural functions than the interactive effects between group and gender. Likewise, we acknowledge that the candidate gene concept in schizophrenia has been largely replaced by genome wide association studies—the former has largely failed to yield insights into the genetic basis of schizophrenia (Collins et al., 2012; Farrell et al., 2015). Moreover, evidence for a direct association between schizophrenia and COMT variation is mixed: a number of meta-analyses have failed to find any such association (Munafò et al., 2005; Okochi et al., 2009), while others demonstrate a clear association (Costas et al., 2011; González-Castro et al., 2016; Williams et al., 2007). Critically, COMT polymorphisms appear to be reliably associated with differential higher order processing (Mier et al., 2010), and appear to be a valid marker of dopaminergic function relative to other candidate genes (Tunbridge et al., 2019). In this context, our present findings of differential effects of COMT variation on neural functions related to higher-order processing in individuals with liability for schizophrenia and bipolar disorder are interesting, though must be interpreted with caution.

The present study documents differential neurophysiological responses to the DS-CPT in individuals with liability for schizophrenia and bipolar disorder. Both relative groups displayed modulated N1 components, though this modulation appears compensatory only in relatives of patients with schizophrenia. Coupled with a larger number of false positive errors to stimuli with contours shared with target stimuli suggests impaired contour detection may reflect the neural consequences of genetic liability for schizophrenia. Deficits in N2 difference waveforms in patients with schizophrenia suggests a disorder specific abnormality related to object recognition. Diminished P3b amplitudes may constitute an endophenotype for schizophrenia building on our previous findings (Sponheim et al., 2006). Finally, we provide novel evidence that COMT variation differentially impacts neural functions in individuals with genetic liability for schizophrenia versus bipolar disorder. Collectively, our findings suggest that aberrant neural responses and not performance on the DS-CPT better differentiate liability for schizophrenia from bipolar disorder, and that fronto-parietal dysfunction may serve as an endophenotype specific to schizophrenia.

## Data Availability

The data that support the findings of this study are available from the corresponding author upon reasonable request.

## Acknowledgments

This work was supported by grants from the Department of Veterans Affairs Clinical Science Research and Development Service (I01CX001843 to SRS), the National Institutes of Mental Health (R01MH112583 to SRS), and the Minnesota Medical Foundation (SMF-2075-99 to SRS); and by the Mental Health Patient Service Line at the Veterans Affairs Medical Center, Minneapolis Minnesota. The content is solely the responsibility of the authors and does not necessarily represent the official views of the Department of Veterans Affairs or the Minnesota Medical Foundation. These funding sources played no role in study design, data collection, analysis, and interpretation, writing this report, or the decision to submit this article for publication.

We thank the EEG research assistants, clinical assessors and diagnosticians for their contributions to data collection and consultation during data analyses.

## Conflict of interest

The authors have no conflicts of interest to disclose.

## Ethical standards

The authors assert that all procedures contributing to this work comply with the ethical standards of the relevant national and institutional committees on human experimentation and with the Helsinki Declaration of 1975, as revised in 2008.

## Notes

### Competing Interest Statement

The authors have declared no competing interest.

